# Diagnosis, triage and response quality of five large language models on emergency medicine vignettes: a blinded comparison

**DOI:** 10.64898/2026.10.21.26366076

**Authors:** Gurinder (Sunny) Johal, Li (Danny) Liang

**Affiliations:** University of British Columbia & Simon Fraser University; University of British Columbia

**Keywords:** artificial intelligence, large language models, emergency medicine, Canadian Triage and Acuity Scale, patient safety

## Abstract

**Objectives:** Emergency care requires appropriate triage and management as well as diagnosis. We compared five large language models on these tasks using the same clinical vignettes.

**Methods:** Seventeen synthetic emergency department vignettes were submitted to Claude Fable 5, GPT-5.6 Sol, Grok 4.5, Gemini 3.6 Flash and Kimi K3 through consumer interfaces on 11–12 August 2026. Two emergency physicians independently assessed the 85 responses with model identities concealed. Outcomes were agreement with reference diagnoses and Canadian Triage and Acuity Scale (CTAS) levels, appropriateness and overall quality (1–5 scales), potential harm and preferred response.

**Results:** Every model correctly named the reference diagnosis in all 17 cases. CTAS exact agreement ranged from 14/17 to 15/17 per model (p=1.0). Of 13 discordant assignments, nine were undertriage and four overtriage. Appropriateness and quality differed across models (both p<0.001): Kimi K3 had the highest mean scores (4.82 and 4.74) and Grok 4.5 the lowest (3.91 and 3.71); Grok 4.5 scored significantly lower than every other model on both scales after adjustment for multiple comparisons. Nine responses were flagged for potential harm: four from Grok 4.5, two each from Gemini 3.6 Flash and GPT-5.6 Sol, one from Claude Fable 5 and none from Kimi K3 (p=0.14).

**Conclusions:** In this selected set of synthetic vignettes, all five models correctly named the reference diagnosis, yet undertriage and potentially harmful omissions occurred and response quality differed between models. These findings do not establish any model’s clinical safety.

**Clinician’s Capsule:** *What is known about the topic?:* Large language models have been evaluated on emergency diagnosis, triage and management, including multi-model comparisons using CTAS.

*What was the study question?:* Do large language model responses that name the correct diagnosis also provide appropriate triage, investigations and management?

*What were the study findings?:* In 17 synthetic vignettes, five models named the reference diagnosis, yet ratings differed and triage errors and potential-harm flags remained.

*Why do these findings matter to clinicians?:* These findings do not establish that any model is safe for clinical use.

## Introduction

Large language models can generate plausible diagnoses and management plans, but evaluations using real patient records have found that they do not reliably follow diagnostic and treatment guidelines.[1] These models have enormous potential as a clinical support tool to improve emergency care, especially in rural and remote areas where residency-trained emergency physicians are often not available.

Emergency medicine studies have compared several models across multiple clinical tasks using simulated cases,[2,3] and triage has been assessed with the Canadian Triage and Acuity Scale (CTAS)[4] in single-model and multi-model vignette studies[5,6] and with the Emergency Severity Index in real emergency department (ED) presentations.[7] However, to the best of our knowledge, there are no existing Canadian studies that evaluate multiple large language models on both CTAS assignment as well as appropriateness of investigation and management recommendations in emergency medicine settings.

We compared five large language models on a common set of emergency vignettes to examine whether responses that name the correct diagnosis also provide appropriate triage, investigations, and management.

## Methods

### Study design and vignettes

We conducted a comparative simulation study with blinded physician assessment. No patients or patient records were involved, and patients and the public were not involved in the design, conduct, reporting or dissemination of this research. The TRIPOD-LLM guideline informed reporting.[8]

We drafted 20 candidate vignettes spanning five clinical domains and CTAS levels 1–3, with assistance from Mistral Le Chat (Think mode), which was not evaluated. Both investigators reviewed each case and its reference answer; three cases without agreement on the reference answer were excluded and four were edited before model testing. Each final case included demographics, history, vital signs and examination findings. Reference answers (diagnosis, acceptable alternatives, CTAS level, investigations, management, disposition and red flags) were finalized before querying and represent the investigators’ clinical judgment.

### Models and querying

We evaluated Claude Fable 5 (Anthropic), GPT-5.6 Sol (OpenAI), Grok 4.5 (xAI), Gemini 3.6 Flash (Google) and Kimi K3 (Moonshot AI), an open-weight model,[9] using the configurations in table 1. Four models were queried on 11 August 2026 and Kimi K3 on 12 August 2026, through consumer web interfaces, with other settings (including web search) at platform defaults, and with account memory and personalization disabled. Each model received all 17 vignettes in one session as a single prompt, in the same order, requesting for each case a most likely diagnosis, two to four differential diagnoses, a CTAS level with justification, initial investigations, management and disposition within 250 words, and instructing that cases be treated independently. One response per case was retained for 85 responses total.

**Table 1.**
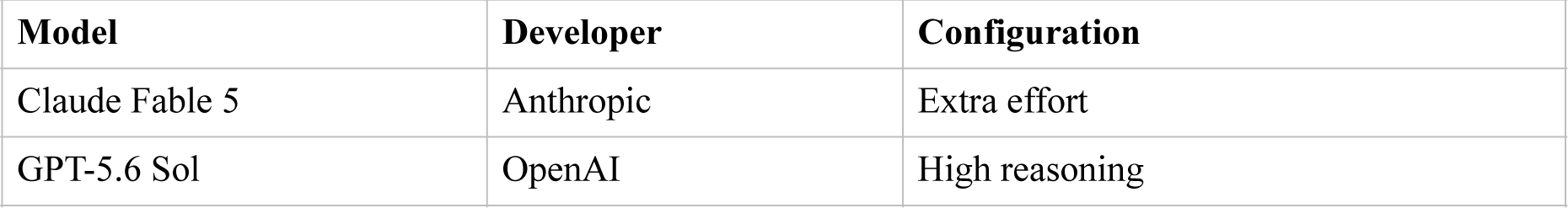
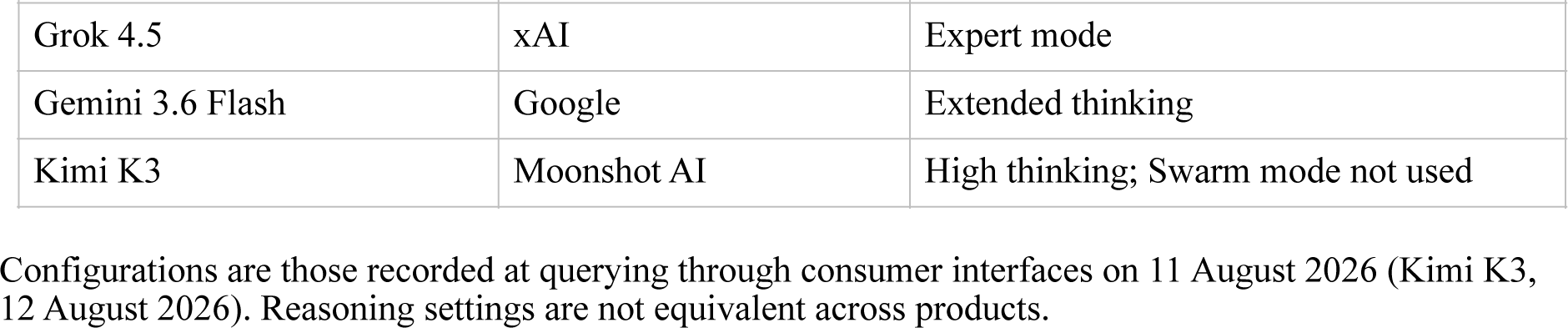
Models and configurations.

### Blinding and outcomes

A script standardized formatting, removed model-identifying text and assigned letters A–E by block randomization, with the model-to-letter mapping varying across vignettes; the decoding key remained sealed until scoring was complete. Both physicians independently assessed every response; one had also collected the outputs.

Diagnosis agreement required the stated most likely diagnosis to match the reference, allowing clinical synonyms. CTAS assignments lower (more urgent) or higher than the reference level were classified as overtriage and undertriage, respectively.

Physicians rated appropriateness of investigations and management (5, all essential steps and nothing incorrect; 4, minor omissions unlikely to change the clinical course; 3, one essential omission or a meaningful non-dangerous error) and overall quality (5, usable without edits, to 1, unusable) on anchored 1–5 scales. Potential harm was a separate binary judgment that an action or omission could plausibly worsen the patient’s outcome if followed, recorded with a rationale; disagreements were resolved by discussion to consensus while both physicians remained blinded to model identity, and other ratings were retained as scored. Each physician selected one preferred response per vignette and could record any perceived model recognition.

### Statistical analysis

The vignette was the matched unit (17 blocks). Binary outcomes are reported with Wilson 95% confidence intervals (CIs) and compared across models with exact Cochran’s Q; overtriage and undertriage comparisons were exploratory. For each response the two physicians’ scores were averaged, and models were compared with Friedman tests followed by two-sided Wilcoxon signed-rank tests (zero differences discarded) with Holm adjustment across the 10 model pairs within each scale; matched-pairs rank-biserial correlations describe effect sizes. Pre-consensus agreement was assessed with Cohen’s kappa for harm and with a two-way random-effects, absolute-agreement intraclass correlation coefficient for the mean of two raters, ICC(2,2), for the rating scales. Preference selections are descriptive. No sample size calculation was performed, and nonsignificant comparisons do not establish equivalence. Analyses used Python 3.12.3, SciPy 1.17.1 and statsmodels 0.14.6; all reported statistics were reproduced with separately written code and in IBM SPSS Statistics.

## Results

### Case mix and rater agreement

The cases comprised four cardiovascular or respiratory, three neurological, four trauma or surgical, three toxicological, environmental or psychiatric, and three pediatric or obstetric vignettes; seven were reference CTAS 1, eight CTAS 2 and two CTAS 3. Raters agreed on diagnosis for all 85 responses. Before consensus, one rater flagged nine responses for potential harm and the other two (agreement 78/85, 91.8%; kappa 0.34); all seven disagreements were resolved towards flagging harm. ICC(2,2) was 0.65 (95% CI 0.31–0.81) for appropriateness and 0.75 (0.62–0.84) for quality. Raters chose the same preferred response in 8/17 vignettes, and neither recorded a model-identification guess.

### Diagnosis and triage

Every model named the reference diagnosis correctly in 17/17 cases (95% CI 81.6%–100%). CTAS exact agreement ranged from 14/17 to 15/17 per model (p=1.0; table 2). Of 13 discordant assignments, nine were undertriage and four overtriage; seven undertriage assignments occurred in two reference CTAS 1 vignettes. Kimi K3 overtriaged three cases and undertriaged none; each other model undertriaged two or three.

**Table 2.** CTAS agreement and potential harm by model (17 responses per model)

| **Model** | **CTAS exact, n (%) [95% CI]** | **Overtriage, n** | **Undertriage, n** | **Potential harm, n (%) [95% CI]** |
|---|---|---|---|---|
| Kimi K3 | 14 (82.4) [59.0–93.8] | 3 | 0 | 0 (0.0) [0.0–18.4] |
| Claude Fable 5 | 14 (82.4) [59.0–93.8] | 0 | 3 | 1 (5.9) [1.0–27.0] |
| Gemini 3.6 Flash | 15 (88.2) [65.7–96.7] | 0 | 2 | 2 (11.8) [3.3–34.3] |
| GPT-5.6 Sol | 14 (82.4) [59.0–93.8] | 1 | 2 | 2 (11.8) [3.3–34.3] |
| Grok 4.5 | 15 (88.2) [65.7–96.7] | 0 | 2 | 4 (23.5) [9.6–47.3] |

All models named the reference diagnosis in 17/17 cases. Overtriage denotes a lower (more urgent) CTAS level than the reference and undertriage a higher number. Harm flags reflect blinded physician consensus. Comparisons across models: exact agreement p=1.0; harm p=0.14; exploratory overtriage p=0.080 and undertriage p=0.30.

### Response ratings and preference

Appropriateness and quality differed across models (both p<0.001; table 3 and figure 1). After adjustment for multiple comparisons, Grok 4.5 scored lower than each other model on both scales (adjusted p 0.004–0.041 for appropriateness and 0.003–0.008 for quality; absolute rank-biserial correlations 0.92–1.00), and Kimi K3 scored higher than Gemini 3.6 Flash on appropriateness (mean difference 0.38; adjusted p=0.046). No other pairwise comparison reached significance. Kimi K3 received 18 of 34 preference selections and Claude Fable 5 received 11.

**Table 3.**
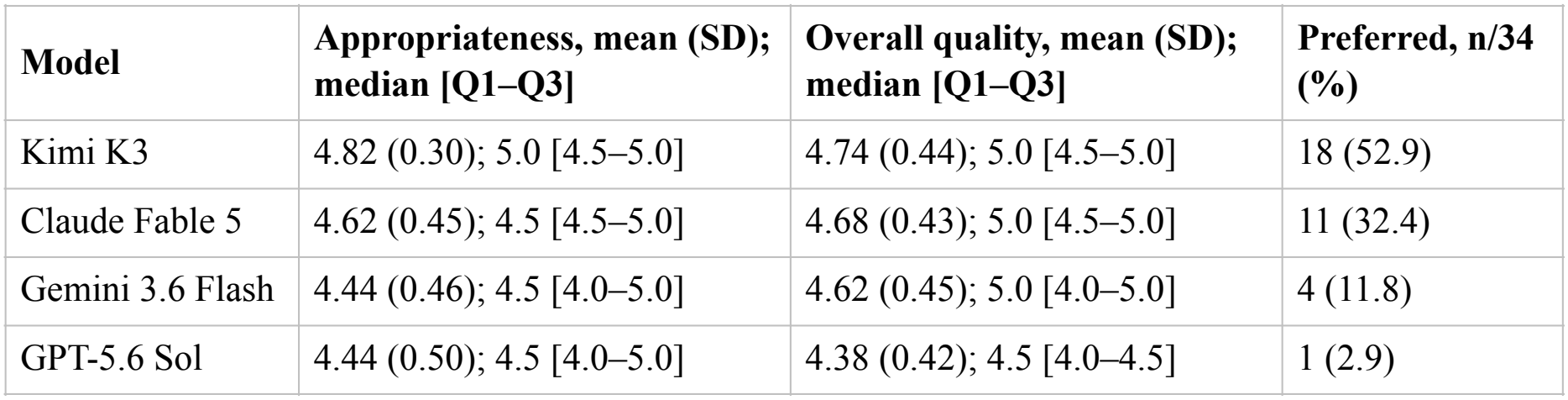
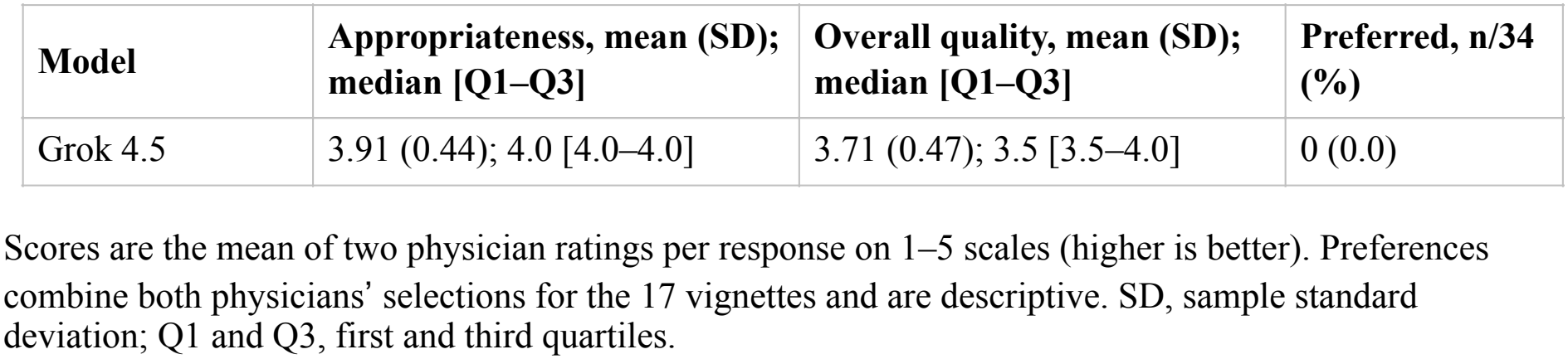
Physician ratings and preferred responses by model (17 responses per model)

**Figure 1.**
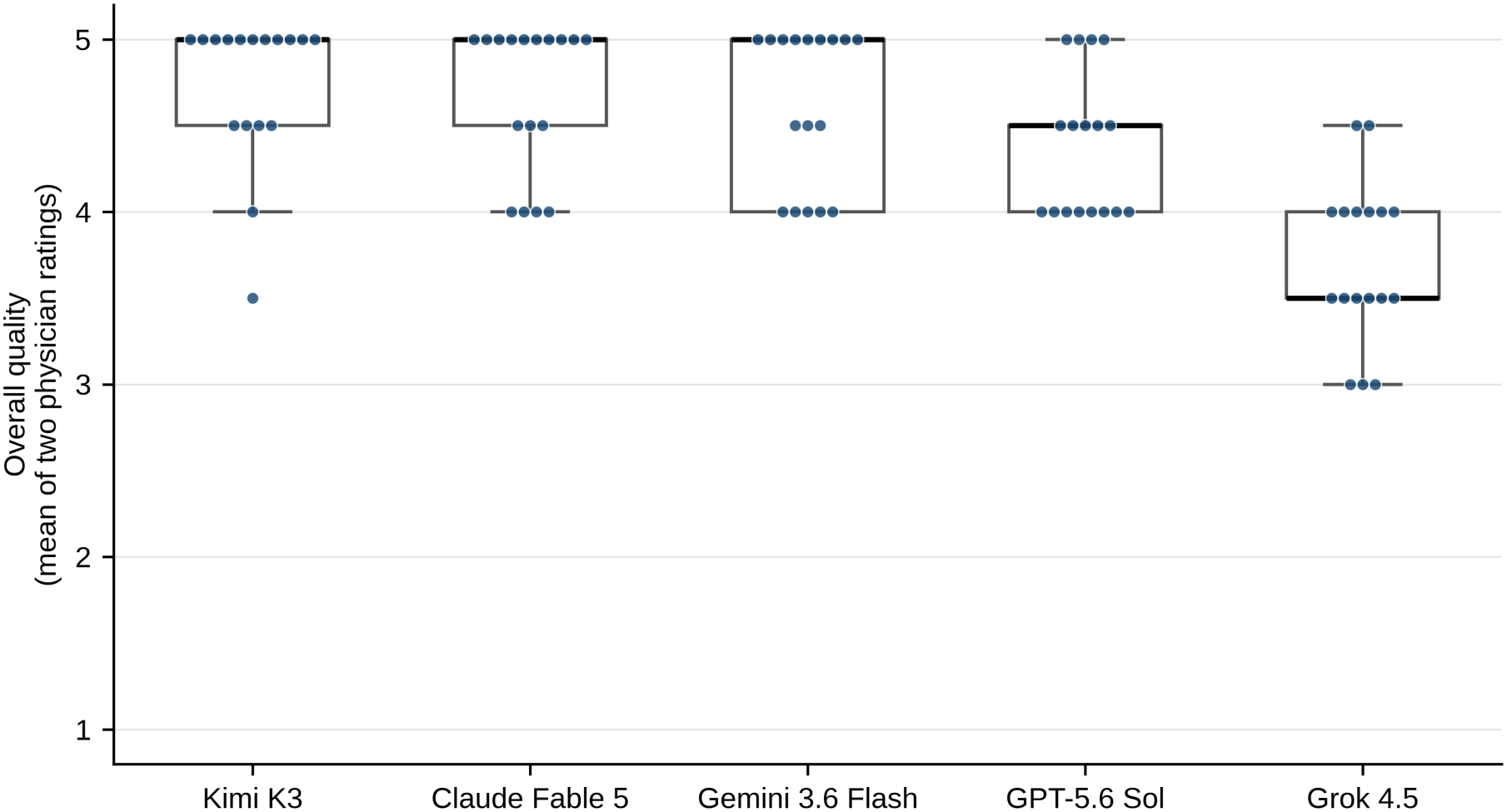
Overall quality ratings by model. Each point is the mean of two physician ratings for one response (17 responses per model). Boxes show the median and first and third quartiles; whiskers extend to the most extreme values within 1.5 times the interquartile range. Scores range from 1 (unusable) to 5 (usable without edits).

### Potential harm

Nine responses (10.6%) were flagged after consensus (table 2; p=0.14). Recorded concerns included omission of blood transfusion in two responses to an ectopic pregnancy, head computed tomography and activated charcoal in three responses to a tricyclic antidepressant overdose, vasopressors in two responses to neonatal sepsis, and RhD status or anti-D immunoglobulin in two responses to a septic abortion at 16 weeks’ gestation. All nine had CTAS assignments matching the reference.

## Discussion

### Interpretation of Findings

All five models named the reference diagnosis in every vignette, but this did not ensure correct triage or complete management. Grok 4.5 scored lower than each other model on both rating scales. Kimi K3 had the highest mean ratings but differed significantly only from Grok 4.5 and, for appropriateness, from Gemini 3.6 Flash. Harm and triage events were too sparse to rank models by safety; zero harm flags among 17 Kimi K3 responses is compatible with a true rate of up to 18.4% (Wilson upper 95% limit).

### Comparison to Previous Studies

Previous multi-model evaluations used different case sets, triage scales and reference standards,[2,3,6,7] so their results cannot be compared directly with ours. The diagnosis ceiling applies only to these selected synthetic cases and does not indicate diagnostic performance in real presentations.

### Strengths and Limitations

To the best of our knowledge, this is the first Canadian study that evaluated large language models in an emergency medicine setting on both triage and appropriateness of investigation and management recommendations.

This study has several important limitations. First, the same investigators wrote the cases, reference answers and rubric and rated the responses; excluding cases without reference-answer agreement may have favoured unambiguous presentations. Second, since all cases were CTAS 1-3, and all but two cases were CTAS 1-2, this study may not accurately reflect the breadth and acuity mix in real-life emergency rooms. Likewise, we cannot infer how the models will perform on CTAS 4-5 cases based on this study. Third, due to our CTAS distribution, since CTAS 1 cases cannot be overtriaged, the acuity mix constrains the direction of triage error, and seven of the nine undertriage assignments came from two vignettes. Fourth, while agreement before consensus was fair, flags combined concerns of differing severity, and harm and appropriateness were not always concordant. Flagged omissions also depend on patient-specific indications, such as airway protection before activated charcoal and RhD status before anti-D immunoglobulin. ICC intervals do not account for clustering within vignettes, and ratings were concentrated at the upper end of both scales (range 3–5), which limits the ICC. Finally, each model answered all cases once, in a single session, within a 250-word limit and through consumer interfaces whose reasoning settings and retrieval defaults differ, and model updates limit reproducibility. With 17 matched blocks and sparse events, inference is limited; larger evaluations with independently developed cases and raters and repeated queries are needed.

### Clinical Implications

Large language models, once clinically validated, have great promise to improve emergency care. Evaluations of these tools should assess management and urgency, not diagnosis alone.

### Research Implications

Future directions include clinical validation of the large language models using real-life clinical cases where accurate presentation histories and past medical conditions are often not readily available, using a larger number of clinical vignettes that include CTAS 4 and 5 cases to better capture the full breadth of emergency medicine, as well as comparing clinical performance on triage, investigations and management with a panel of emergency physicians.

### Conclusion

Large language models are some of the most promising tools to improve emergency care, especially in rural and remote settings. In 17 synthetic ED vignettes, five large language models consistently named the reference diagnosis but differed in physician-rated quality, and triage errors and potentially harmful omissions persisted. Future studies on the clinical validation of large language models using a wide range of real-life cases, with physician performance as the benchmark, are needed. Evaluations should assess management and urgency alongside diagnosis; these findings do not establish comparative clinical safety.

## Data Availability

All data produced in the present study are available upon reasonable request to the authors.

## Acknowledgements

We thank Samar Hejazi, PhD, MN, Research & Implementation Scientist, Department of Evaluation and Research Services, Fraser Health, for independently reproducing the statistical analyses.

## Disclosures

### Ethics approval

Not applicable. The study used synthetic cases only and involved no human participants or patient data; the physician raters were the investigators.

### Patient consent for publication

Not applicable.

### Use of artificial intelligence

Mistral Le Chat (Think mode), which was not among the evaluated models, assisted with drafting candidate vignettes. Claude (Anthropic) supported study planning, development of the blinding script and scoring workbooks, and verification of reported statistics. Grok, Mistral and ChatGPT/Codex each generated analysis code following a prespecified plan. ChatGPT/Codex, Claude and Grok also assisted with manuscript editing, literature review and reference checking. Physicians finalized the reference answers and performed all scoring. The authors take responsibility for the manuscript’s content.

